# Expression of HER2/neu in premalignant lesions of oral cavity

**DOI:** 10.1101/2020.07.24.20161489

**Authors:** Vansh Verma, Ashesh Kumar Jha, Karsing Patiri, Nikhil Arora

## Abstract

**Context:** Molecular alterations in premalignant lesions of oral cavity are not well known, many reports and have found increased HER2 expression to be correlated with poor prognosis in oral cancer. However, literature on expression of HER2 in premalignant lesions is limited and data is conflicting in nature. Overexpression of HER2 in premalignant lesions may denote its positive contribution in malignant transformation of these lesions.

**Aims:** To evaluate the expression of HER2 in premalignant lesions of oral cavity.

**Settings and Design:** In this prospective observational study of 2 months, patients attending OPD at Department of ENT and meeting the inclusion criteria were included.

**Methods and Material:** 23 samples of Leukoplakia and 1 sample of oral lichen planus were stained by routine H&E to confirm clinical diagnosis and assess dysplasia if any, 5 samples of normal mucosa were used as control. Immunohistochemical staining for HER2 was done. ASCO/CAP 2018 guidelines were used for reporting the results.

**Statistical analysis used:** Percentage of lesions expressing cytoplasmic or membranous expression was calculated.

**Results:** 1 sample of Leukoplakia with severe dysplasia expressed focal membranous staining. 20% leukoplakia lesions expressed cytoplasmic staining. Staining was not observed in oral lichen planus and leucoplakia without dysplasia.

**Conclusions:** Membranous expression in Severe dysplasia and higher expression in oral cancer is in concordance with the multistep theory of carcinogenesis. Larger studies are needed if HER2 is to be proposed as a marker for oral premalignant lesions. Significance of cytoplasmic staining in oral premalignant lesions needs to be elucidated.

**Key Messages:** To the best of our knowledge, this is the first report of focal membranous expression of HER2 in leucoplakia in India. 20% leucoplakia with dysplasia expressed cytoplasmic staining. The significance of cytoplasmic staining needs to be further explored.

## Introduction

Oral cancer is one of the most frequently encountered malignancy in this part of the world, accounting for approximately 30% of all the malignancies in India.^[1]^ Most of these are squamous cell carcinoma and are preceded by premalignant lesions, and approximately 60% of oral cancers appear as white keratotic lesions.^[2,3]^ Despite the recent advancement in medicine, the overall prognosis and 5-year mortality rates of Oral squamous cell carcinoma have not changed drastically due to delayed presentation, field cancerization and occurrence of second primary tumour. Moreover, therapeutic interventions such as surgery, chemotherapy or radiotherapy for potentially curable diseases can increase the morbidity and affect the quality of life.^[4]^

Premalignant lesions are considered as precursors of invasive carcinoma and are frequently reported in the oral cavity. The overall prevalence of precancerous lesions in India has been reported to be as high as 8.4%.^[5]^ In India, prevalence of Leukoplakia varies from 0.2 to 5.2% making it the most commonly encountered lesion, The risk of malignant transformation of these lesions is variable, ranging from 0.3% to 10% depending upon the grade of dysplasia, location, clinical type and other changes at the molecular level^[6]^. Identification of high-risk premalignant lesions and appropriate interventions at early stage are desirable to ensure a favourable outcome.

Hence it seems prudent to look for molecular changes in premalignant lesions to predict their malignant potential.

HER2/neu (c-erbB-2 oncogene) is located on chromosome 17 and is a member of the erbB family of receptors. Upon ligand binding, the receptors have potential to homodimerize or heterodimerize with other members of the ErbB protein family, resulting in autophosphorylation of their tyrosine kinase domains and downstream signaling.^[7]^ These proteins are expressed in most epithelial cell layers and have a key role in development.^[8]^ Currently, cetuximab, a monoclonal antibody to EGFR, is the only FDA-approved molecular targeting agent for the treatment of primary or metastatic head and neck squamous cell carcinoma. HER2/neu shares sequence homology to EGFR, which is overexpressed in oral squamous cell carcinoma, oral premalignant and dysplastic lesions of oral cavity.^[9,10]^ Her2/neu acts through RAS-MAP Kinase pathway and plays a central role in cell proliferation and survival. It inhibits cell death through the mTOR pathway. It is well known that HER2/neu is found to be significant in maintaining oncogenesis in several malignancies, notably breast, ovary, stomach, pancreas, kidney and others.^[11]^ A sequential increase in her2 expression starting at dysplastic stage has been reported in stomach, gallbladder, breast and other malignancies.^[12-14]^ A gradual increase in EGFR and HER2 expression starting at precancerous stages in hamster cheek mouse system for oral carcinogenesis has also been reported.^[15]^

The PI3K pathway is implicated in premalignant lesions’ pathogenesis, and HER2 acts via the same leading to increased cellular proliferation and survival.^[16,17]^ EGFR is found to be upregulated in premalignant lesions, expression of Her-2 could possibly implicate formation of EGFR&HER2 heterodimers further upregulating the PI3K pathway.

Reported Expression levels in Oral squamous cell carcinoma have been diverse, ranging from 0 to 47%.^[18-21]^ Some reports have found Her-2 expression to be linked with prognosis in oral squamous cell carcinoma, Xia et al used IHC in 80 patients with HSCC and considered cytoplasmic and membranous staining for Her-2 as positive and reported Her-2 expression to be the most significant factor in predicting disease outcome, Cavalot at al. concluded that Her2/neu expression and lymph nodal status are independent predictors of disease free survival in head and neck squamous cell carcinoma patients.^[17,20]^

Previous studies (Table 1) done regarding the expression of Her2 in premalignant lesions of oral cavity have shown a variable expression ranging from 0 to as high as 80% in severe dysplasia. Fong et al in Taiwan reported 25% premalignant samples expressing Her-2 using immunohistochemistry but did not divide the staining pattern into membranous or cytoplasmic.^[22]^ Seifi et al in Iran reported 11% premalignant samples expressing membranous Her-2, on the other hand Werkmeister et al. reported chromosome 17 deletions in approximately 15% lesions.^[23,24]^ Rautava et al. reported 35% premalignant samples showing slightly increased staining and 13% lesions with markedly increased staining, somewhat consistent with Werkmeister et al.^[25]^ Kobayashi et al. in Japan took only non dysplastic leukoplakias and found no samples with increased expression, this was contrary to Wilkman et al. who found increased HER2 expression in some samples with epithelial hyperplasia and dysplasia, they reported 57% premalignant lesions positive for Her-2.^[26,27]^ Li Hou et also considered cytoplasmic HER2 expression as positive and reported 13,71,80% samples with mild, moderate and severe dysplasia respectively expressing Her-2.^[28]^

**Table 1.** Previously done studies on HER2 expression in oral premalignant lesions.

| Serial number | author | Method | HER2 expression |
| --- | --- | --- | --- |
| 1. | Fong et al | IHC | 25% |
| 2. | Seifi et al | IHC | 11% |
| 3. | Rautava et al | IHC | 35% moderately increased staining, 13% markedly increased |
| 4. | Werkmeister et al | PCR | 15% samples showed chromosome 17 deletions in Non-dysplastic Leukoplakia |
| 5. | Wilkman et al | IHC | 57% |
| 6. | Mandal et al | IHC | Cytoplasmic staining in a few |
| 7. | Kumar et al | IHC | Cytoplasmic staining in a few |
| 8. | Kobayashi et al | IHC&PCR | Cytoplasmic staining and high levels of HER2 mRNA levels but no significant increase |
| 9. | Hou et al | IHC | 13%,71%,80% positivity in mild,moderate and severe dysplasia respectively |

The key molecular players associated with oral premalignant lesions are TIMP1, Bcl-2, TGF-Beta, HER2 interacts with the abovementioned either in a binary manner or via signal transducers; thus HER2 is a potential candidate gene for pathogenesis of oral premalignant lesions.^[29,30]^ Since oral malignancies have shown HER2 expression, overexpression of the same in premalignant lesions may denote its positive contribution in malignant transformation of these lesions. Available literature shows conflicting and divergent results concerning overexpression of HER 2/neu in such lesions. Hence this study was undertaken to evaluate the expression of HER2 in premalignant lesions of oral cavity.Our aims and objectives are as follows-

1. Expression of HER2/neu in premalignant lesions of the oral cavity.
2. Correlation of HER2/neu expression with different types of premalignant lesions

## Materials and Methods

This prospective observational study was conducted over a period of 2 months in the Department of Surgery, Otolaryngology and Pathology. During this period all consenting patients attending OPD at Department of Surgery and ENT meeting the inclusion criteria were included. Ethical clearance was obtained from the Institution ethics committee.

- Inclusion criteria- All diagnosed cases of oral premalignant lesions
- Exclusion criteria – Patients with inflammatory and Malignant lesions of oral cavity

Biopsy taken from the premalignant lesions of the study subject after obtaing informed consent. This biopsy sample was processed in the Department of Pathology. Control was taken from the adjacent healthy mucosal tissue upon patient’s consent.

In **routine H&E staining**, dewaxed sections in xylene were taken through graded alcohol and brought to water, staining with harris haematoxylin for 8 minutes was done, then washed until bluing occurred, afterwards they were differentiated in 1% acid alcohol for 5 seconds and washed underwater again until bluing occurred, then staining with Y eosin was done for 2 minutes, washed with distilled water, again dehydration was done in graded alcohol solutions, passed through xylene i.e. a clearing agent, finally mounting was done with DPX. Histological diagnosis was made to confirm the clinical diagnosis.

Dysplasia, if the present was be graded according to the WHO 2017 system for grading of dysplasia. Hyperplasia is also considered under mild dysplasia as per this classification.^[31]^

- Mild dysplasia: Architectural changes limited to the lower third of the epithelium along with the cytological atypia.
- Moderate dysplasia: Architectural changes extending to the middle third of the epithelium.
- Severe dysplasia/carcinoma in situ: abnormal maturation extending from basal cells to a level above the midpoint of epithelium to entire thickness of epithelium.

Further, unstained sections were taken for immunohistochemical staining.

**Immunohistochemical-expression** of HER2/neu was determined on 4-micron thick paraffin sections; all steps were carried out in moist and humid container.

1. Slides were marked.
2. The 4-micron sections were deparaffinised by putting them on a hot plate and dipping in xylene.
3. They were hydrated with graded ethanol, followed by being brought to water.
4. The sections were placed in 3% hydrogen peroxide in methanol for 30 minutes.
5. A coplin jar filled with 10Mm Citrate buffer (pH-6) and covered with a lid was preheated in a pressure cooker for 5 minutes.
6. The slides were put into the buffer, covered with a clinging film and heat in the microwave for 10 minutes at low power to allow antigen retrieval. This allows antibody binding to epitope, which gets masked during formalin fixation.
7. The slides along with buffer were allowed to cool down.
8. Three washes with TRIS buffer were given.
9. The nonspecific proteins were blocked with 5% milk block.
10. Three washes with TRIS buffer were given.
11. The tissue was incubated with primary antibody at 4 degree Celsius overnight.
12. Three washes with TRIS buffer were given.
13. Then secondary antibody was added for 30 minutes.
14. TRIS wash was given thrice.
15. Then tertiary antibody was added for another 30 minutes.
16. Three washes with PBS buffer were given.
17. DAB was applied on all slides and reaction will was monitored under a microscope.
18. The slides were then immersed in distilled water as soon as crisp brown staining was be seen.
19. The slides were counterstained with H&E stain.
20. The slides were dehydrated in a graded alcohol solution.
21. They were passed through xylene, which acts as a clearing agent.
22. Finally, the slides were mounted with DPX.

The staining pattern was recorded as – membranous or cytoplasmic.

The results were recorded as per CAP/ASCO 2018 guidelines.^[32]^ (Table 2).

**Table 2.**
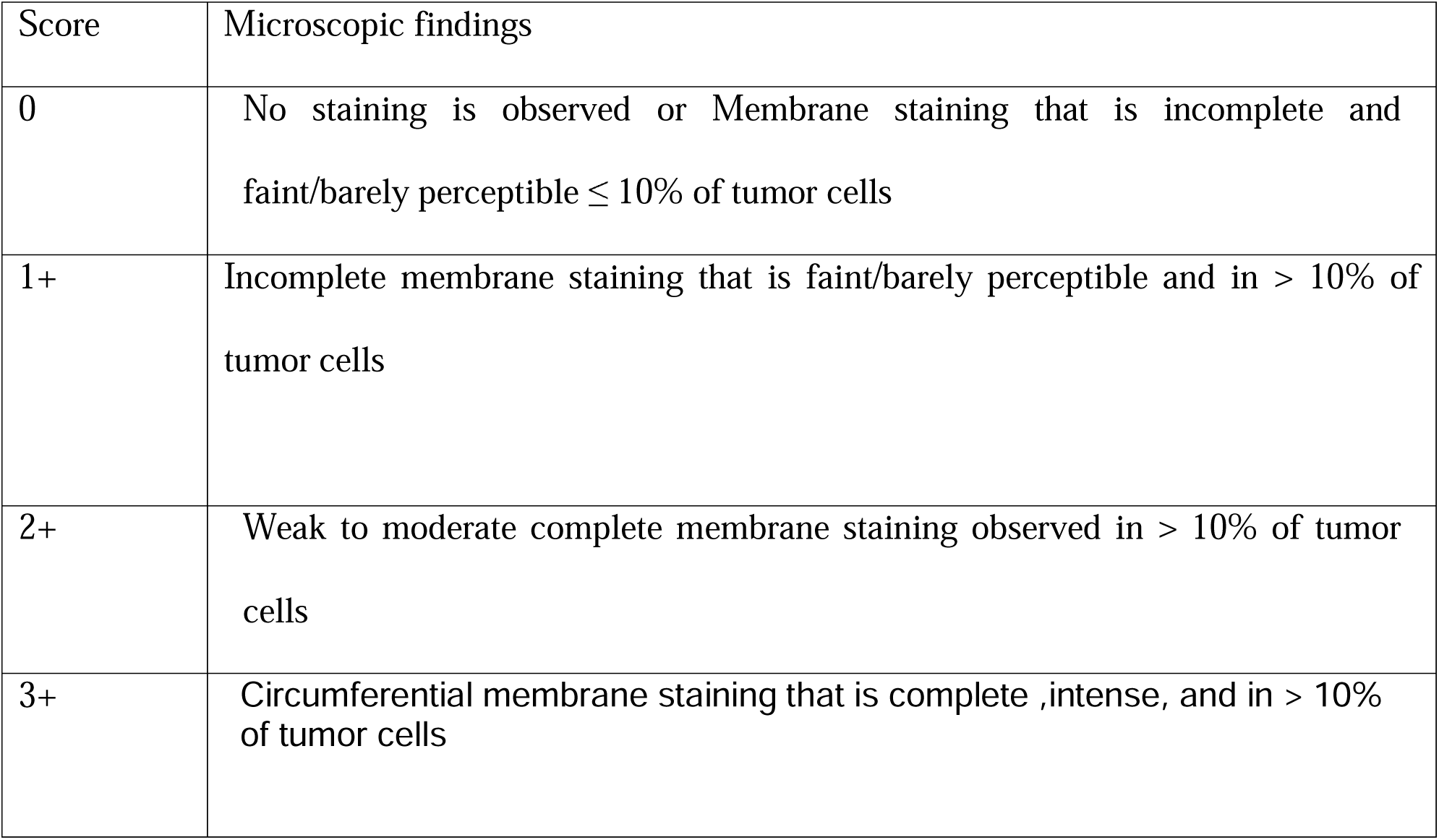
ASCO/CAP 2018 guidelines.

The percentage of positive cells were determined along with intensity; the results were tabulated.

The results from IHC were analysed; patients were classified depending upon the type of premalignant lesion. Afterwards statistical analysis was done.

### Statistical analysis

Percentage of lesions expressing HER2 was calculated using Excel.

## RESULTS

Out of the 24 patients taken for this study, 22 were males, and 2 were females, male to female ratio was 11:1. Age range observed is 28 to 60 years, average age of the patients is 43 years.

All patients in this study were tobacco users.

Clinical characterization-23 patients in the study were characterized as having a white patch, whereas 1 patient was characterized having Oral Lichen Planus(Table 3).

**Table 3.**
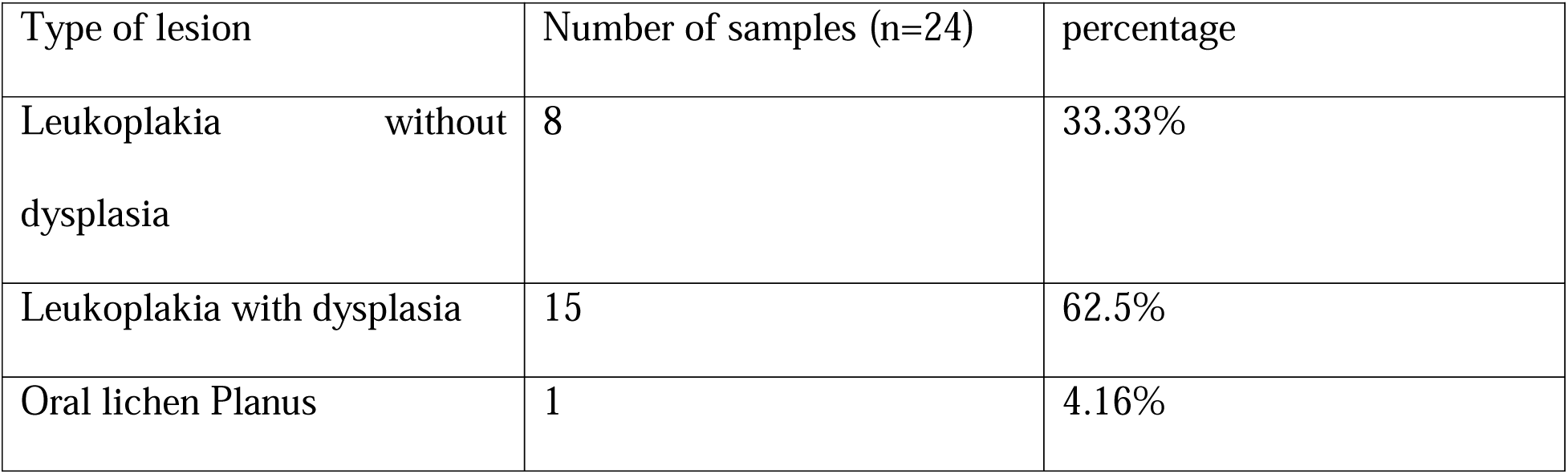
Various premalignant lesions encountered in the study subjects.

Site distribution of lesions-of the 23 lesions, 17 were located on the buccal mucosa, 5 were located on the tongue and one each was located on lower lip and gingiva(Table4).

**Table 4.**
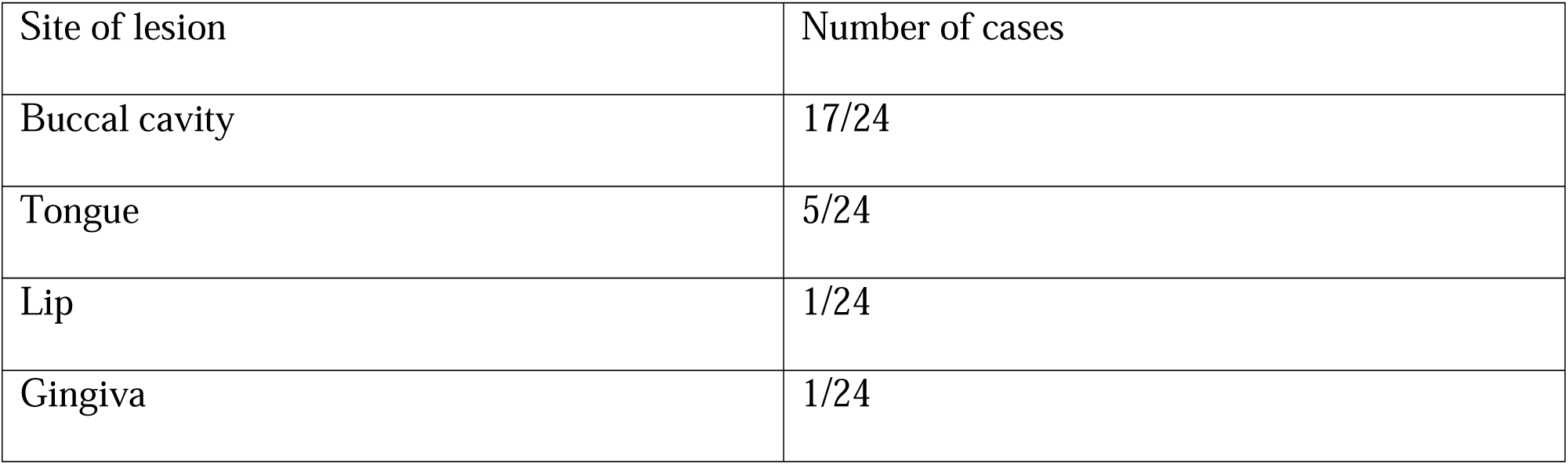
site of premalignant lesions.

**Table 5.**
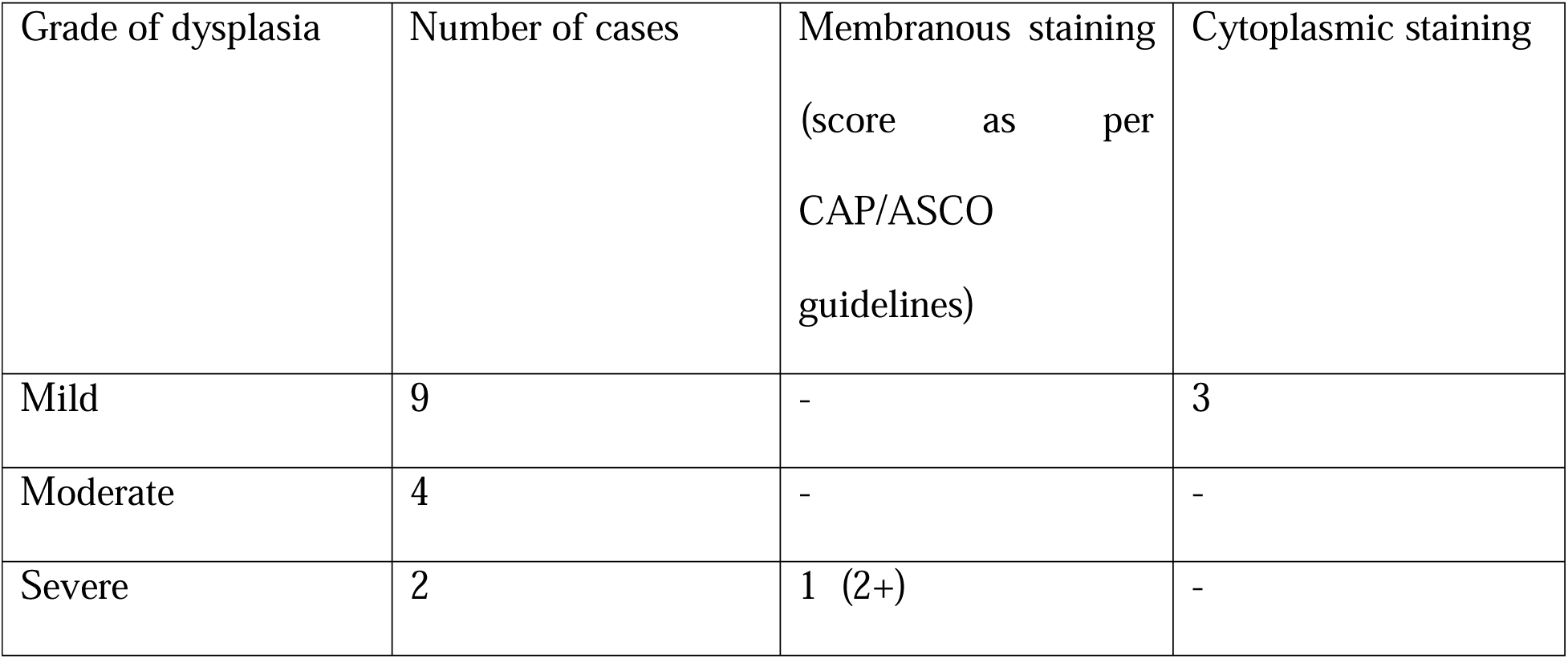
staining characteristics of premalignant lesions.

**Table 6.**
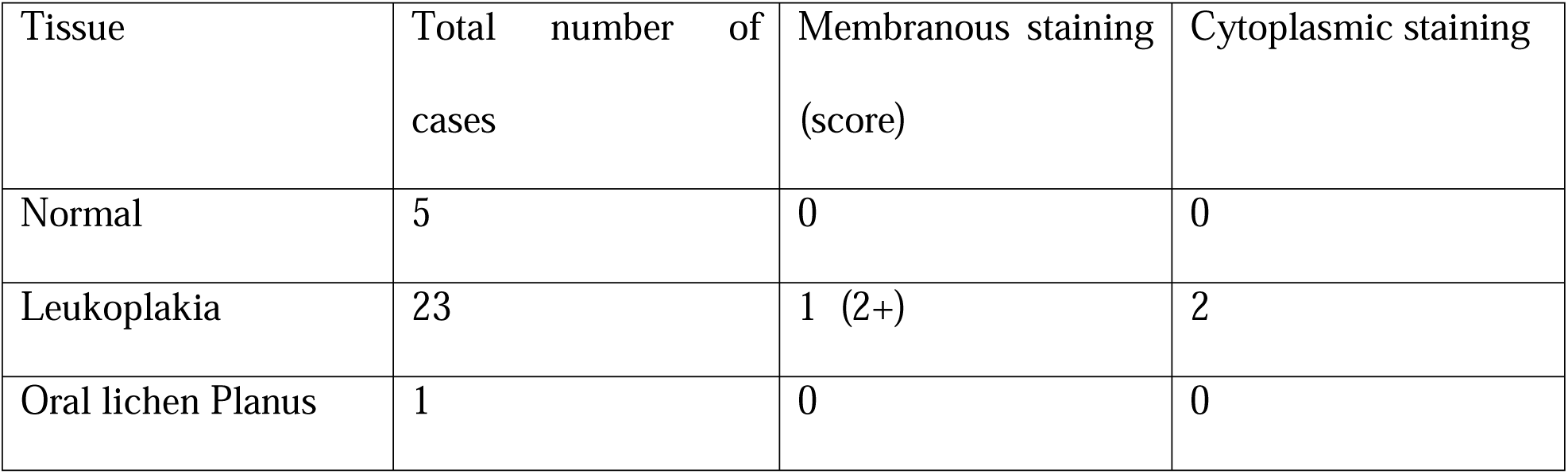
staining pattern with respect to type of lesion.

Of the 23 histologically proven Leukoplakia cases, 9 cases had dysplasia and 6 cases had hyperplasia, which is now considered under mild dysplasia as per WHO 2017 criteria.^[31]^ 65.21 % samples of leukoplakia samples had dysplasia.

HER2 staining can be observed in membranous/cytoplasmic fashion, However 2+ is considered as equivocal &3+ is considered positive as per the ASCO/CAP guidelines.

Focal membranous positivity (2+) (Figure 1) was seen in one case of leukoplakia with severe dysplasia i.e. 4.34% of Leukoplakia samples in this study.

**Figure 1.**
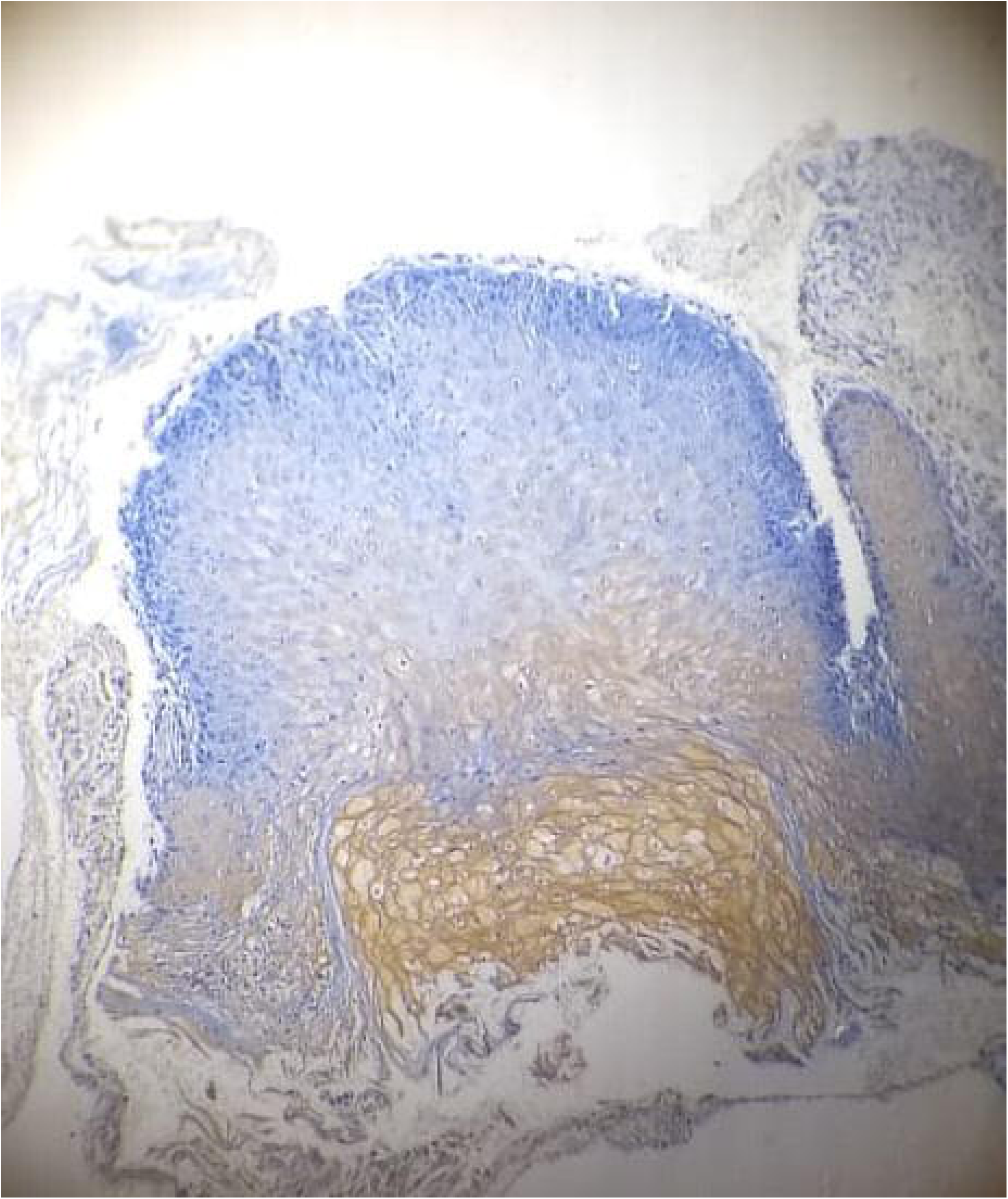
Focal Membranous staining (2+) for HER2 in leukoplakia sample with severe dysplasia.

Cytoplasmic staining was observed in three cases of Leukoplakia with mild dysplasia i.e. 12.5% of all lesions in this study and 13.04% of leukoplakia in this study. In cases of leukoplakia with dysplasia, 20% lesions expressed cytoplasmic staining. Membranous/cytoplasmic HER2 expression was not noted in any sample without dysplasia.

## DISCUSSION

On analysis of the demographic composition, males outnumbered the females, this is in concordance with higher tobacco consumption in men according to social and cultural norms. Tobacco consumption has been regarded as the primary etiological factor for development of premalignant lesions, all patients in this study were tobacco users. Buccal mucosa was the most common site accounting for 70.8% of cases, followed by 20.8% of cases in the tongue, and 4% cases each in lip and gingiva.

HER2 staining can be observed as membranous or/and cytoplasmic, however ASCO/CAP 2018 guidelines consider only membranous expression (2+&3+) as positive staining for Breast cancer.^[32]^ Fluorescent in situ hybridization is recommended for 2+ cases as per CAP/ASCO 2018 guidelines.

In our study, one out of 23 leukoplakia samples demonstrated focal membranous and cytoplasmic staining and 3 samples demonstrated cytoplasmic staining.

Taking into account the dysplastic samples, 1 out of 2 i.e. 50% samples of severe dysplasia expressed focal membranous with cytoplasmic staining and 3 mildly dysplastic lesions expressed cytoplasmic expression, no sample of moderate dysplasia stained for HER2. HER2 staining in oral premalignant lesions has been reported to be between 0 to 80% in the existing literature.(Table 1)

Oral lichen planus is not known to express HER2 and our findings are consistent with the existing literature.^[26]^ We report 4.34% leukoplakia expressing focal membranous with cytoplasmic positivity, our expression rate is lower as compared to studies elsewhere, Seifi et al reported 11% samples having membranous positivity and Wilkman et al reported 60% samples having membranous staining.^[23,27]^ For studies which did not divide staining pattern as membranous or cytoplasmic, such comparison is not possible. Our findings are in contrast to the previously done 2 studies in India which did not report membranous staining, they did not quantify percentage of samples showing cytoplasmic staining.^[33,34]^ However, we report 13.04% leukoplakia samples having cytoplasmic staining.

Fong et al and Rautava et al considered both membranous and cytoplasmic staining as positive and reported 25% & 35% samples showing increased HER2 expression.^[26,29]^ If we consider both cytoplasmic and membranous staining, we get 4/24 samples showing increased expression i.e. 16.6%.

With regards to dysplasia, we report 1/9 samples expressing membranous positivity, our findings are in contrast to wilkman et al who reported HER2 membranous expression in 57% dysplasia.^[27]^ Lin Hou et al considered cytoplasmic staining as positive and reported 13%,71% and 80% samples expressing HER2 in mild, moderate and severe dysplasia respectively. In our study, we did not find any sample with moderate dysplasia expressing HER2. Recent studies have reported less HER2 positivity as compared to the ones done previously, this could be possibly due to differences in methods and specificity of the antibody used.

Similarly in oral squamous cell carcinoma, various studies have reported expression between 0 to 47%.^[18-21]^ The major difficulty in comparing immunohistochemical studies is the variety of antibodies, even procedural variations like incubation time, antigen retrieval and lack of standardization in scoring may cause variation.

Controversial results in different studies may be due to

- Using different IHC methods:direct or indirect
- Using different types of antibody
- Lack of standardization in reporting results
- Location of lesions within the oral cavity
- Significance of cytoplasmic HER2

Significance of cytoplasmic staining is a widely debated topic, in colorectal and oral cancer, few studies have shown cytoplasmic association to be linked with prognosis, indicating a role in pathogenesis.^[17,20]^ A plausible explanation could be homodimerization of cytoplasmic domains of HER2 leading to tyrosine kinase activation and PI3K overexpression further regulating cell proliferation.^[35]^

Polysomy of chromosome 17 is a well known step in oral carcinogenesis, a study reported that 15.3% lesions with pure cytoplasmic staining had chromosome 17 polysomy, whereas all samples with moderate membranous with cytoplasmic staining had chromosome 17 polysomy, even in non HER2 amplified breast cancer, HER2 immunohistochemical expression has been found to be linked with polysomy of chromosome 17. It has been suggested that this genetic aberration may result in a significant increase of Her-2 gene copies in the tumor cells and an increased Her-2 protein production to the level that could be demonstrated by immunohistochemistry as overexpressed.^[36]^ In Breast cancer it was suggested that cytoplasmic HER2 could be a different or a truncated 155 kD protein as compared to the 185 kD protein expressed on the membrane, however in colorectal cancer osako et al reported 185 kD protein in both the fractions and reported no 155 kD peptide, indicating an activated intracellular HER2 receptor. Some authors suggest that cytoplasmic HER2 expression is an artifact or occurs due to cross reactivity possibly with keratin, others propose that it may reflect true protein expression and incomplete receptor degradation.

For other types of cancer, it has been reported that HER2 overexpression can occur via mechanisms other than gene amplification, such as via increased levels of promoter-binding proteins as in gastric cancer where elevated levels of binding proteins to the TATA-box were observed, which led to HER2 overexpression. Theoretically, mutations in downstream targets of HER2, for example KRAS, might also influence the expression of HER2 via affected feedback processes. These mechanisms could play a role in the observed cytoplasmic HER2 overexpression without any gene amplification. However, their significance in oral cancers is uncertain.^[36]^

HER2 expression can have significant impact on pathogenesis of premalignant lesions and oral cancer, EGFR is known to be expressed in oral premalignant lesions, HER2 can form heterodimers with EGFR and activate tyrosine kinase pathway leading to PI3K overexpression.

Trastuzumab targets only membranous HER2 receptors, however lapatinib, a relatively newer drug recently approved for breast cancer patients targets intracellular tyrosine kinase and may possibly be used for premalignant/malignant lesions showing cytoplasmic HER2 staining. For completely unraveling the role of HER2 in oral premalgnant and malignant lesions, follow up studies with treatment and simultaneous measurement of HER2 levels will be needed.Moreover no clear cut guidelines for HER2 immunohistochemical expression reporting exist, this is in contrast to breast, gastric cancer where clear cut guidelines exist. This leads to irregularities in reporting results.

## Data Availability

The authors confirm that the data supporting the findings of this study are available within the article.

## LEGEND

6. Table 6- Staining characteristics of dysplastic samples

## REFERENCES

1. Sharma S, Satyanarayana L, Asthana S, Shivalingesh K, Goutham BS, Ramachandra S. Oral cancer statistics in India on the basis of first report of 29 population-based cancer registries. J Oral Maxillofac Pathol. 2018; 22:18–26.

2. Warnakulasuriya S, Johnson NW, van der Waal I. Nomenclature and classification of potentially malignant disorders of the oral mucosa. J Oral Pathol Med. 2007;36:575–580.

3. Ali AA, Al-Sharabi AK, Aguirre JM, Nahas R. A study of 342 oral keratotic white lesions induced by qat chewing among 2500 Yemeni. J Oral Pathol Med 2004;33:368–72.

4. Jadhav KB, Gupta N. Clinicopathological prognostic implicators of oral squamous cell carcinoma: need to understand and revise. N Am J Med Sci. 2013;5:671–679.

5. Byakodi R, Shipurkar A, Byakodi S, Marathe K. Prevalence of oral soft tissue lesions in Sangli, India. J Community Health. 2011;36:756–759.

6. Kulkarni MR. Head and neck cancer burden in India. Int J Head Neck Surg. 2013;4:29– 35.

7. Dittrich A, Gautrey H, Browell D, Tyson-Capper A. The HER2 Signaling Network in Breast Cancer--Like a Spider in its Web. J Mammary Gland Biol Neoplasia. 2014;19:253–270.

8. Arkhipov A, Shan Y, Kim ET, Dror RO, Shaw DE. Her2 activation mechanism reflects evolutionary preservation of asymmetric ectodomain dimers in the human EGFR family. Elife. 2013;2:e00708.

9. Zhang H, Liu J, Fu X, Yang A. Identification of Key Genes and Pathways in Tongue Squamous Cell Carcinoma Using Bioinformatics Analysis. Med Sci Monit. 2017;23:5924–5932

10. Rajeswari M, Saraswathi T. Expression of epithelial growth factor receptor in oral epithelial dysplastic lesions. Journal of Oral and Maxillofacial Pathology. 2012;16:183.

11. Iqbal N, Iqbal N. Human epidermal growth factor receptor 2 (HER2) in cancers:overexpression and therapeutic implications. Mol Biol Int. 2014;2014:852748.

12. Ieni A, Barresi V, Rigoli L, Caruso R, Tuccari G. HER2 Status in Premalignant, Early, and Advanced Neoplastic Lesions of the Stomach. Disease Markers. 2015;2015:1–10.

13. Barreto S, Dutt A, Chaudhary A. A genetic model for gallbladder carcinogenesis and its dissemination. Annals of Oncology. 2014;25:1086–1097.

14. Kader T, Hill P, Rakha EA, Campbell IG, Gorringe KL. Atypical ductal hyperplasia: update on diagnosis, management, and molecular landscape. Breast Cancer Res. 2018;20:39.

15. Vairaktaris E, Moulavassili P, Loukeri S, Spyridonidou S, Yapijakis C, Derka S, et al. Abundance and localization of skeletal muscle-related erbB2 may stimulate tumour growth during initial stages of oral oncogenesis. J Musculoskelet Neuronal Interact 2007;7:185–90.

16. Martins F, de Sousa SC, Dos Santos E, Woo SB, Gallottini M. PI3K-AKT-mTOR pathway proteins are differently expressed in oral carcinogenesis. J. Oral. Pathol. Med. 2016;45:746–752.

17. Cavalot A, Martone T, Roggero N, Brondino G, Pagano M, Cortesina G. Prognostic impact of HER-2/neu expression on squamous head and neck carcinomas. Head Neck. 2007;29:655–64.

18. Sardari Y, Pardis S, Tadbir AA, Ashraf MJ, Fattahi MJ, Ebrahimi H, et al. HER2/neu expression in head and neck squamous cell carcinoma patients is not significantly elevated. Asian Pac J Cancer Prev. 2012;13:2891–6.

19. Vats S, Ganesh M S, Agarwal A. Human epidermal growth factor receptor 2 neu expression in head and neck squamous cell cancers and its clinicopathological correlation: Results from an Indian cancer center. Indian J Pathol Microbiol 2018;61:313–8.

20. Xia W, Lau YK, Zhang HZ, Liu AR, Li L, Kiyokawa N, et al. Strong correlation between c-erbB-2 overexpression and overall survival of patients with oral squamous cell carcinoma. Clin Cancer Res 1997;3:3–9.

21. Khan AJ, King BL, Smith BD, Smith GL, DiGiovanna MP, Carter D, et al. Characterization of the HER-2/neu oncogene by immunohistochemical and fluorescence in situ hybridization analysis in oral and oropharyngeal squamous cell carcinoma. Clin Cancer Res 2002;8:540–8.

22. Fong Y, Chou S, Hung K, Wu H, Kao S. An Investigation of the Differential Expression of Her2/neu Gene Expression in Normal Oral Mucosa, Epithelial Dysplasia, and Oral Squamous Cell Carcinoma in Taiwan. Journal of the Chinese Medical Association. 2008;71:123–127.

23. Seifi S, Shafaei SN, Nosrati K, Ariaeifar B. Lack of elevated HER2/neu expression in epithelial dysplasia and oral squamous cell carcinoma in Iran. Asian Pac J Cancer Prev. 2009;10:661–4.

24. Werkmeister R, Brandt B, Joos U. Aberrations of erbB-1 and erbB-2 oncogenes in non-dysplastic leukoplakias of the oral cavity. British Journal of Oral and Maxillofacial Surgery. 1999;37:477–480.

25. Rautava J, Jee KJ, Miettinen PJ, Nagy B, Myllykangas S, Odell EW, et al. ERBB receptors in developing, dysplastic and malignant oral epithelia. Oral Oncol. 2008;44:227–35.

26. Kobayashi H, Kumagai K, Gotoh A, Eguchi T, Yamada H, Hamada Y, Suzuki S, Suzuki R. Upregulation of epidermal growth factor receptor 4 in oral leukoplakia. Int J Oral Sci. 2013;5:14–20.

27. Wilkman TS, Hietanen JH, Malmstrom MJ, Konttinen YT. Immunohistochemical analysis of the oncoprotein c-erbB-2 expression in oral benign and malignant lesions. Int J Oral Maxillofac Surg. 1998;27:209–212.

28. Hou L., Shi D., Tu S.M., Zhang H.Z., Hung M.C., Ling D. Oral cancer progression and c-erbB-2/neu proto-oncogene expression. Cancer Lett. 1992;65:215–220.

29. Tsai HP, Chen SC, Chien HT, Jan YY, Chao TC, Chen MF, Hsieh LL. Relationships between serum HER2 ECD, TIMP-1 and clinical outcomes in Taiwanese breast cancer. World J Surg Oncol.2012;10:42.

30. Carpenter RL, Han W, Paw I, Lo HW. HER2 phosphorylates and destabilizes pro-apoptotic PUMA, leading to antagonized apoptosis in cancer cells. PLoS One. 2013;8:e78836.

31. Müller S. Update from the 4th Edition of the World Health Organization of Head and Neck Tumours: Tumours of the Oral Cavity and Mobile Tongue. Head and Neck Pathology. 2017;11:33–40.

32. Wolff A, Hammond M, Allison K, Harvey B, Mangu P, Bartlett J et al. Human Epidermal Growth Factor Receptor 2 Testing in Breast Cancer: American Society of Clinical Oncology/College of American Pathologists Clinical Practice Guideline Focused Update. Journal of Clinical Oncology. 2018;36:2105–2122.

33. Singla S, Singla G, Zaheer S, Rawat DS, Mandal AK. Expression of p53, epidermal growth factor receptor, c-erbB2 in oral leukoplakias and oral squamous cell carcinomas. J Can Res Ther. 2018;14:388–93.

34. Kumar P, Kane S, Rathod GP. Coexpression of p53 and Ki 67 and lack of c-erbB2 expression in oral leukoplakias in India. Braz Oral Res 2012;26:228–34.

35. Blok EJ, et al. Cytoplasmic overexpression of HER2: a key factor in colorectal Cancer. Clin Med Insights Oncol. 2013;7:41–51.

36. Papavasileiou D, Tosios K, Christopoulos P, Goutas N, Vlachodimitropoulos D. Her-2 immunohistochemical expression in oral squamous cell carcinomas is associated with polysomy of chromosome 17, not Her-2 amplification. Head Neck Pathol. 2009;3:263– 70.

